# The effect of calcium and vitamin D on bone mineral density in patients taking glucocorticoid therapies: Protocol for a systematic review and network meta-analysis

**DOI:** 10.1101/2020.05.24.20112177

**Authors:** Jiawen Deng

## Abstract

Glucocorticoid (GC) administration is an effective therapy commonly used in the treatment of autoimmune and inflammatory diseases. However, the use of GC can give rise to serious complications. The main detrimental side effect of GC therapy is significant bone loss, resulting in glucocorticoid-induced osteoporosis (GIOP).

We performed a systematic review and network meta-analysis (NMA) to evaluate whether the use of calcium supplementation, with or without vitamin D, vitamin D metabolites and vitamin D analogues is capable of increasing bone mineral density (BMD) at the lumbar spine, femoral neck, and hip in adult patients undergoing glucocorticoid therapies compared to no treatment.

## INTRODUCTION

Glucocorticoid (GC) is a class of steroids commonly prescribed to patients suffering from chronic inflammatory diseases. Its excellent ability of inhibiting a variety of inflammatory mediators makes it an ideal choice for treating autoimmune diseases, such as systemic lupus erythematosus, or chronic respiratory diseases such as asthma[1,2]. A study conducted by Gotzsche & Johansen had found that, when used for treating conditions such as rheumatoid arthritis and post-operative inflammation, glucocorticoids can yield better outcomes compared to NSAIDs[3–5]. Because of glucocorticoids’ superiority over other anti-inflammatory therapies, long-term glucocorticoid use is prevalent in clinical settings. In the first national estimate of oral GC usage in the US, it was reported that over 2,000,000 patients had taken GCs from 1999-2008. The mean duration of GC use was over 1,000 days, with 28% of patients having used glucocorticoids for over 5 years[6].

While the efficacy of glucocorticoids surpasses their non-steroidal counterparts, their side effects are also much greater. One particularly worrying aspect about the chronic use of GC is its destructive effect on bones. Glucocorticoids inhibit factors required for osteoblast proliferation and differentiation, resulting in the inhibition of bone formation, causing osteoporosis[7–9]. GIOP occurs in 30-50% of patients placed on long-term GC therapy, and the risk of fracture increases significantly during the first 3-6 months of glucocorticoid use[8,10].

The American College of Rheumatology (ACR) has published guidelines supporting the use of vitamin D and calcium in adult patients taking glucocorticoids, with or without an additional, concurrent antiosteoporotic therapy[11]. However, ACR’s latest guidelines did not make any specific recommendations as to the use of vitamin D metabolites and/or vitamin D analogs. Our previous investigations had revealed that certain vitamin D metabolites and analogs, namingly alfacalcidol and calcitriol, may have greater anti-fracture efficacy compared to plain vitamin D_3_ (i.e. cholecalciferol). Therefore, we propose a systematic review and network meta-analysis to compare the effect of calcium supplementation, vitamin D, D analogs or D metabolites on the bone mineral density of patients undergoing glucocorticoid therapies compared to no treatment.

### Methods

This research protocol will be written in accordance with the Preferred Reporting Items for Systematic Reviews and Meta-Analyses (PRISMA) protocol[12]. This study is currently being registered on PROSPERO.

Our electronic literature search will be conducted to address our research question phrased as follows in the PICO framework:

1. **Population:** The study population consists of adult patients undergoing active glucocorticoid therapy at baseline, or adult patients who are about to undergo glucocorticoid therapy during the course of the RCT study.
2. **Interventions:** Interventions include calcium supplementations (e.g. calcium carbonate, calcium lactate, calcium gluconate, etc.), plain vitamin D (e.g. cholecalciferol, ergocalciferol, etc.), and vitamin D analogs/metabolites (e.g. alfacalcidol, calcitriol, etc.).
3. **Comparator** The efficacy of the interventions will be compared to placebo and untreated (no administration of any antiosteoporotic therapy, calcium or vitamin D supplementations).
4. **Outcome** Our primary outcomes of interest include percentage change of bone mineral density from baseline at the following sites: 1) lumbar spine, femoral neck, and total hip.

Only data from randomized controlled trials will be included for analysis as RCT data is the highest in the hierarchy of evidence in terms of different trial designs[13–15]. We will include academic publications of all languages in our analysis.

### Electronic Database Search

We will conduct a librarian-assisted search of MEDLINE, EMBASE, PubMed, Web of Science, CINAHL and the Cochrane Library using search strategies developed to address our PICO question. Major Chinese databases, including Wanfang Data, Wanfang Med Online, CNKI, and VIP will also be searched using a custom Chinese search strategy. In addition, clinicaltrials.gov and the WHO International Clinical Trials Registry Platform will be hand searched for unpublished research data.

We will utilize the same search strategy as our previous study regarding GIOP, registered on PROSPERO (ID CRD42019127073). Please see https://doi.org/10.1101/19010520 for sample Chinese and English search strategies.

### Study Selection

All search results yielded by the literature search will undergo two screening processes in duplicate. A PRISMA flowchart will be generated to show the number of inclusions and exclusions at every step of the search and screening process.

We will perform blinded abstract screening in duplicate using Rayyan QCRI, and all conflicts will be resolved via consensus. All results from the literature search, with the exception of duplicate entries, will be uploaded to Rayyan for screening. The search results will be deduplicated using EndNote (X9).

If we encounter promising entries that are in languages other than English or Chinese, we will attempt to recruit local translators to obtain a translated version of the publication. If a local translator is not available, the study will be excluded from the analysis. For entries in Chinese, two reviewers who are fluent in Chinese will perform the screening in duplicate.

The full text screening will be completed in duplicate using a standardized screening checklist, and conflicts resolved via consensus. Papers written in languages other than English or Chinese were translated by a local translators prior to screening. If a study included GC patients but did not report GC specific data as a subgroup, we contacted the authors/data holders to request aggregate level outcome data.

### Data Abstraction

We will extract data from all included trials in duplicate into standardized electronic data extraction sheets created a priori for analysis.

Data extracted include:

**Study Design** - study ID (author name and year of publication), clinical trial ID, study design (e.g. double blind, cross over, double dummy), method for random sequence generation, method for allocation concealment, active arm, comparator, adjuvant therapy, GC dosage during study, GC duration during study, followup period, BMD measurement methodology, authors’ conflict of interest statements.

**Baseline** - # patient randomized, # patient analyzed, # patients lost to followup, median age and range, gender distribution, % postmenupausal, GC dosage at baseline (equivalent to prednisone), GC indication, GC duration at baseline, bone mineral density reading at baseline (in g/cm^2^), % patients who had already experienced at least one fracture at baseline.

**Outcome Related** - Final total hip, femoral neck, and lumbar spine BMD reading at the end of the latest follow up period (g/cm^2^), baseline total hip, femoral neck, and lumbar spine BMD reading (g/cm^2^), percentage and absolute change (in g/cm^2^) in BMD at total hip, femoral neck and lumbar spine regions.

For all continuous outcomes, the standard deviation will be extracted to provide an estimate of variance in the study population. For RCTs that failed to report measures of variance (i.e. standard deviation) the 95% confidence interval or p value will be used to derive the standard deviation based on methods recommended by the Cochrane Handbook[16]. If a publication published neither the standard deviation nor the confidence interval, we will estimate the standard deviation for percentage changes based on the reported mean and variance of baseline and final BMD readings using methods described here: https://www.ncbi.nlm.nih.gov/books/NBK33484/. If a study did not report the mean percentage change, we will calculate the percentage change using the baseline and final mean BMD readings.

### Data Synthesis

All statistical analyses will be conducted using R. We will perform random effects model network meta-analyses using the gemtc library which is based on the Bayesian probability framework.

For each outcome, we will generate a network diagram. A network diagram illustrates treatment arms as “nodes”, and the trials that provide direct comparison between arms as “edges”. The size of the nodes represent the total number of direct comparisons that include the represented treatment arm in the network, while the thickness of the edges represent the number of publications that offer a direct connection between the connected nodes.

Results of the NMA will be reported as mean differences (MD) for all outcomes. Forest plots will be generated to illustrate treatment efficacies and rankings graphically. We will also generate SUCRA (surface under the cumulative ranking curve) scores, which is a numerical representation of the likelihood that a given treatment will rank first in a specific outcome. A SUCRA score closer to 1 represents a higher likelihood of ranking first and thus a greater chance of being the best.

Global inconsistency within the network will be measured using the I^2^ value, which represents the amount of study heterogeneity[17]. A high measure of global inconsistency suggests that the included studies report conflicting efficacies; an I^2^ value closer to 0% indicates a more coherent network.

### Risk of Bias

The quality of the included studies will be evaluated using The Cochrane Collaboration’s Tool for Assessing Risk of Bias in duplicate[18]. We will use “low”, “unclear”, and “high” to evaluate the effectiveness of the methodologies used for random sequence generation, allocation concealment, blinding of participants and personnel, blinding of outcome assessment, as well as for the completeness of outcome data, the presence of selective reporting, and other biases for each study. The same standards will be applied to studies of all languages.

## GRADE

While our risk of bias analysis will be used to evaluate the quality of individual studies, the overall quality of evidence within networks will be assessed using the Grading of Recommendations Assessment, Development, and Evaluation (GRADE) framework in duplicate for each outcome[19]. In this network meta-analysis, we will use CINeMA, an online network quality appraisal tool developed in accordance to the GRADE framework to evaluate and synthesize the quality of direct and indirect evidence for each outcome network, according to the latest methodological guidelines. CINeMA evaluates network quality based on six criteria: within-study bias (risk of bias), across-study bias, indirectness, imprecision, heterogeneity, and incoherence[20]. We will report the results of our GRADE analysis in conjunction with the statistical significance of treatment efficacies compared to placebo using a summary of findings table.

### Meta-Regression

There are several potential factors for the development of osteoporosis apart from GC therapy, such as gender, post-menopausal status, and age[21,22]. Therefore, meta-regression analyses will be run to check for covariate effects. Meta-regression will be ran on % female in the patient population, % post-menopausal in the patient population, and the median age of the population. We will also run meta-regressions on median cumulative GC dose at baseline and follow up durations.

## Data Availability

No data available for this study.

